# Correlation analysis of latent hepatitis B virus infection gene mutation in unpaid blood donors

**DOI:** 10.1101/2024.07.15.24310447

**Authors:** Wei Yu, Fang Wang, Qiang Liu

**Affiliations:** Jiangxi Province Blood Center, Nanchang City 330052, Jiangxi, China

**Keywords:** HBV, OBI, S, HBsAg

## Abstract

**Objective:** To analyze the HBV S region gene mutation in serum samples of occult hepatitis B virus (HBV) infection (OB1), and to further analyze the mechanism of gene mutation and OBI.

**Methods:** Blood samples from unpaid blood donors from January 2022 to December 2023 were collected and analyzed for serology, liver function indicators, viral load and gene mutation.

**Results:** A total of 90 OBI cases were detected, the detection rate was 0.051%, and the incidence of OBI was correlated with age. The sequence amplification of s region in the blood samples showed the presence of C type and B type genes, both of which occurred in the hydrophilic region.

**Conclusion:** There is no abnormal liver function between OBI and hepatitis B population, and there is no difference in viral load between different serum modes. Mutations in the HBVS region are more likely to cause OBI, which provides a scientific basis for more sensitive HBV detection.

## Introduction

With the development of society, hepatitis B virus (HBV) is a global health problem. According to the latest report, there are 200 million chronic hepatitis B patients in the world, and hepatitis B patients will further progress to cirrhosis, liver failure, and even liver cancer. The number of deaths due to chronic hepatitis B is about 1 million every year, and it is increasing year by year [1, 2]. At present, the method of detecting hepatitis B is to detect serological markers, of which hepatitis B surface antigen (HBsAg) is a crucial serological indicator. However, some hepatitis B patients have been found to be negative for HBsAg, which makes the diagnosis of some hepatitis B difficult. Patients who are negative for hepatitis B surface antigen (HBsAg) have occulthepatitis B virus infection (OBI). According to the working group of the European Association for the Study of Liver Diseases in 2008, OBI means that the human body is HBsAg negative and the amount of HBV-DNA virus is less than 200U/ml [3]. It is known that OBI patients generally have low immunity, frequent blood transfusion and frequent vascular puncture [4-6], so it can be inferred that OBI is mainly transmitted through blood transfusion, organ transplantation and other ways.

The research on OBI has lasted for more than 30 years, but the pathogenesis of OBI has not been fully understood at present, which brings great challenges to the clinical and epidemiological studies of OBI. According to previous studies, HBV gene mutation, immunosuppression and insufficient hepatitis B detection technology are the main reasons [7,8]. From the perspective of gene mutation, HBV is more prone to gene invariability, and the mutation region is mainly the envelope protein S gene region, core promoter [9], and the core hydrophilic region MHR of the S gene region is the most important mutation site [10,11]. The mutation frequency in this region is high, which will lead to the binding of antibody and antigen during HBsAg detection. Thus, HBsAg negative results are formed, which has an impact on clinical diagnosis [12].

This study selected 90 OBI patients from January 2022 to December 2023, and explored the epidemiological characteristics of these patients by analyzing the demographic data of these patients. In this study, the HBV genotyping of OBI patients was studied by PCR technology, and the gene status of S region was analyzed. In order to understand OBI infection and genetic mutation characteristics of unpaid blood donors. Therefore, a more sensitive detection strategy is provided for the current HBV detection technology, and the missing detection situation of OBI is constructed, so as to better prevent OBI and diagnose and treat OBI.

## Materials and methods

### Patient population

Serum from unpaid blood donors from January 2022 to December 2023 was collected and OBI patients were screened. The blood samples of blood donors were screened for HBsAg by ELISA reagent (Yingke Xiamen Technology Co., LTD.). A total of 175583 blood samples were collected. Blood samples with negative HBsAg were selected for HBV DNA detection (using Shanghai Bori Biology Department). OBI patients should meet the diagnostic criteria: HBsAg negative and HBVDNA positive after Elisa and NAT detection of HBVDNA. After screening, a total of 90 people met OBI standards. Serum of 10 HBsAg positive and HBV DNA positive patients with HBV infection were selected as positive control.

This trial has been approved by the Ethics Committee.

## Methods

### Liver function index detection

Liver function index including alanine aminotransferase ALT, aspartate aminotransferase AST and total bilirubin were detected in blood samples of OBI group and positive control group to analyze the epidemiology of OBI patients.

### Serological index detection

Blood samples of OBI patients were tested for serological indexes, in which HBsAg and anti-HBS were quantitatively detected by electrochemical luminescence analyzer, and anti-HBE and anti-HBC were detected by ELISA reagent.

### HBV DNA viral load detection

The viral load of OBI blood samples was measured and HBV nucleic acid quantitative detection kit (PCR-fluorescent probe method) was used. The lower limit of detection was 5IU/mL..

#### Analysis of HBV s region gene mutation

Nested PCR was used to amplify the genes in OBI and HBVS region of the positive control group, and the amplified products were sequenced [13]. The first round amplification system: the total volume was 25 uL, including template DNA 10μL, the amount of polymerase was 0.75U, the final concentration of Mg2+ was 2.5mmol/L, the final concentration of primer was 200 nmol /L, and the final concentration of dNTPs was 200 umol /L. The second round of amplification system: the total volume was 50u, containing 10u of the first round of amplification products, the amount of polymerase was 1.5U, and the rest was the same as the first round of amplification. When S1 primer was used for nested PCR, the selected polymerase was TaKaRaLATaq HS. When S2/S3 primers were used for nested PCR, the selected polymerase was TaKaRaEXTaq HS. The first amplification parameters of S2/S3 primers were: 94° C 4 min, 94° C 30s, 56° C 30s, 72° C 30s, a total of 35 cycles, extending at 72° C for 10min; The second round of amplification parameters only increased the annealing temperature to 58□, and the rest were the same as the first round of amplification. The amplification parameters of S1 primer were the same as those of S2 /S3 primer except that the annealing time was extended to 45s and the extension time was extended to 1 min. The second round PCR product 5uL was mixed with 1uL of SYBR Green dye buffer and 1.1% agarose gel electrophoresis was performed. The PCR products with clear amplification bands were sequenced, and the sequencing work was completed by Shanghai Shenggong Biological Engineering Co., LTD

### amplified sequences

The amplified sequenceswere checked by ChromasPro software. The 24 genotype sequences of NCBI HBVA-H8 were compared and analyzed by MEGA6 software. The nucleic acid and amino acid sequences of the s region were compared with previous Chinese HBV reference sequences [14]

### Statistical analysis

SPSS 23.0 was used for statistical analysis of the experimental data. Counting results use case (%) analysis, chi-square test; The measurement results were tested by non-parametric test. If P is< 0.05, there is a statistical difference.

## Results

### Epidemiological characteristics

In this study, a total of 90 OB1 cases were screened from 175583 blood donations, and the infection rate was 0.051%. According to the analysis of demographic information, there was no significant difference between genders (P>0.05), and the difference between ages was statistically significant. The higher the age, the higher the incidence of OBI.

**Table 1.**
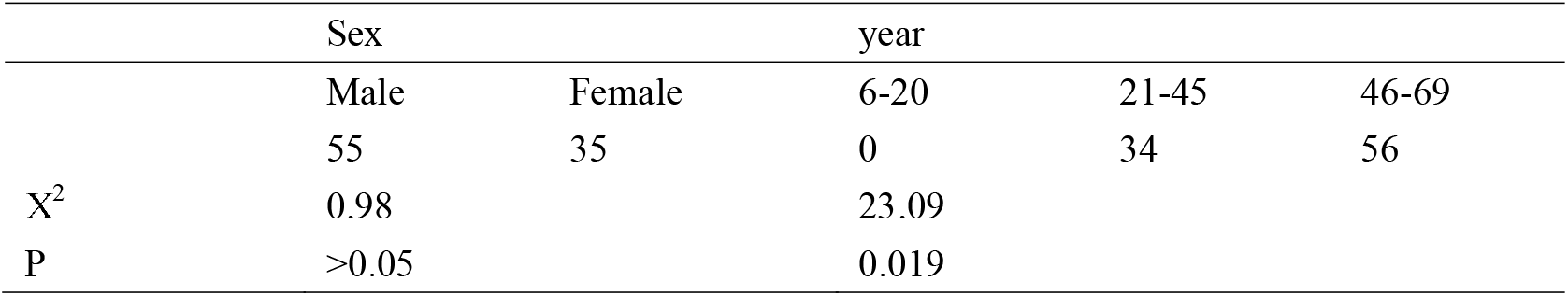
Demographic characteristics of patients.

#### Liver function index

There were no significant differences in ALT, AST and bilirubin between OBI group and positive control group (P>0.05).

**Table 2.** Comparison of liver function indexes between OBI group and positive control group.

|  | OBI | positive control g | P |
| --- | --- | --- | --- |
| ALT ( U/L ) | 10.98±1.43 | 11.23±3.09 | >0.05 |
| AST ( U/L ) | 15.75±2.87 | 14.98±2.98 | >0.05 |
| bilirubin ( μmol/L) | 6.23±1.32 | 6.15±0.98 | >0.05 |

#### Serological indicators

Serological analysis of 90 OBI blood samples showed that 5 types were detected, among which anti-HBS +, anti-HBE +, anti-HBC + accounted for the highest (26.67%), and anti-HBS + accounted for the lowest (11.11%).

**Figure 1.**
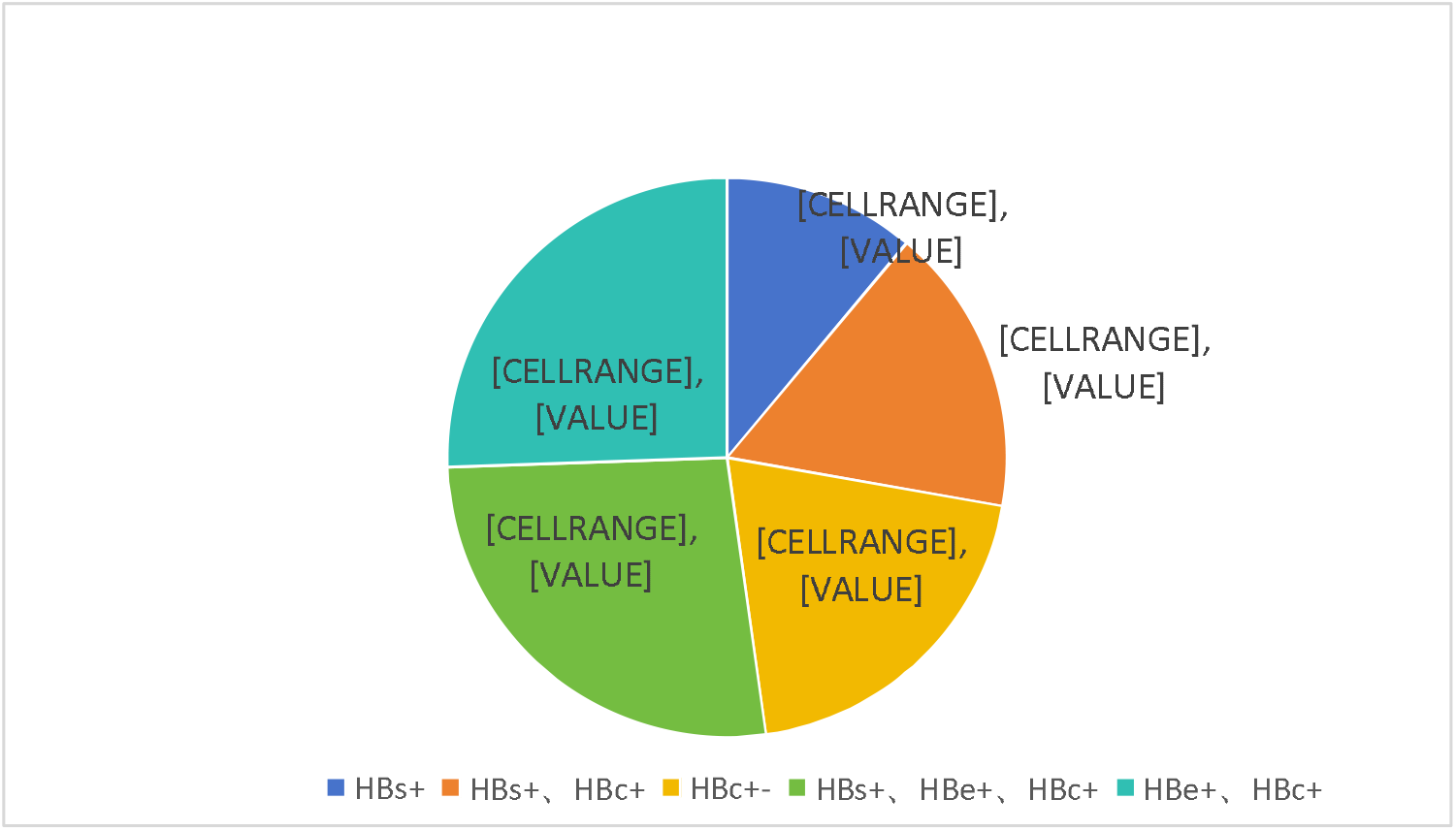
Serological index analysis.

### HBV DNA viral load analysis

different serological antibody types showed little difference in viral load level, and there was no statistical significance between serological type and viral load level (P>0.05). +

**Table 3.**
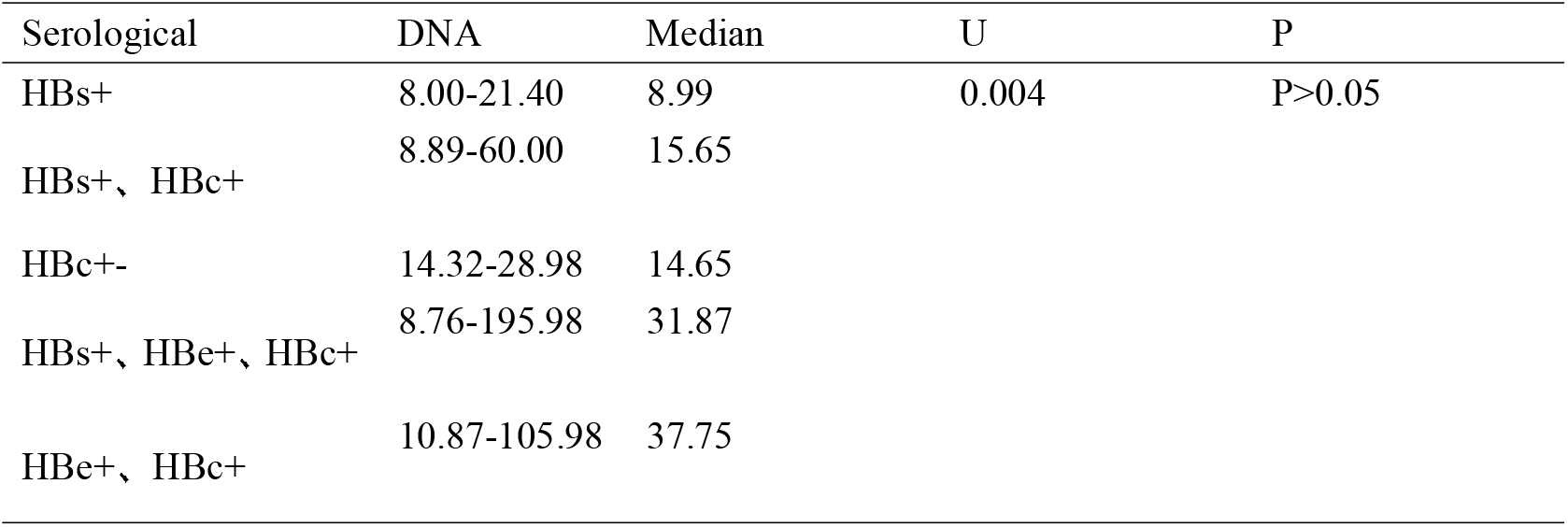
Correlation between serological type and viral load level.

### OBI S Gene mutation analysis

In 90 OBI cases, the S-region gene mutation was analyzed by nested PCR, and 70 cases were sequenced successfully. The genotypes were B type and C type, with 50 cases of B type and 20 cases of C type. A total of 560 gene mutations occurred, including 334 gene mutations in B genotype and 226 gene mutations in C genotype, all of which occurred amino acid mutations in the hydrophilic region.

**Table 4.**
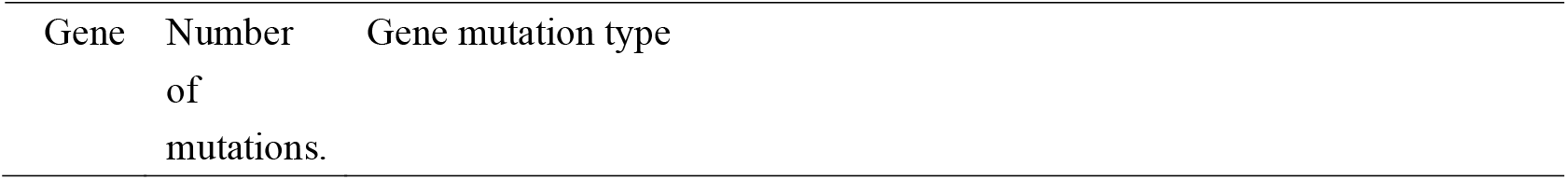
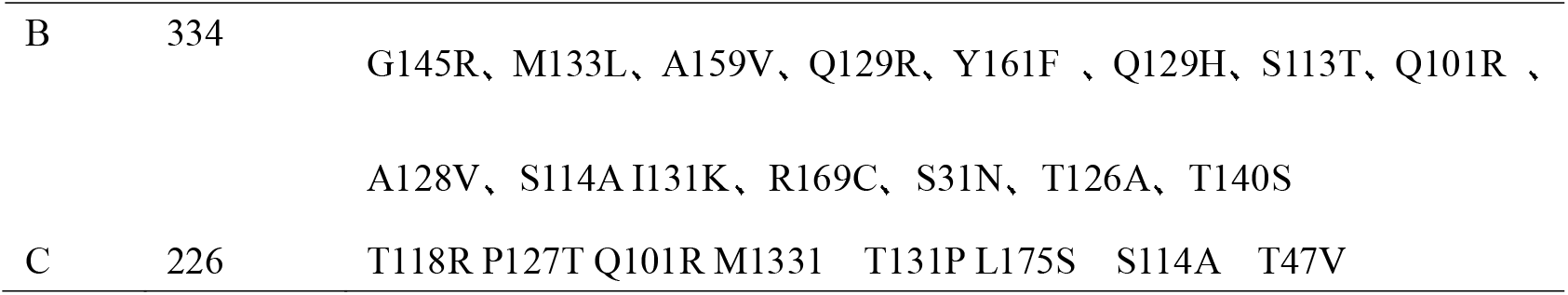
OBI S Gene mutation.

## Discussion

At present, it is believed that the main form of infection of HBV is blood transmission, and HBV has been used as an important screening index in clinical practice, so as to reduce the unsafe situation in the blood transfusion. However, there are limitations in the detection of HBV screening index HBsAg at present, which may lead to missed diagnosis of OBI, thus posing challenges to the safety of blood transfusion [15].

This study found that the incidence of OBI in blood donors was0.051%, which was slightly higher than previously reported, which may be caused by regional differences or limited number of studies [16]. This study found that the incidence of OBI in 46-69 years old population was higher than that in other age groups, accounting for 62.2%, and there was no correlation between gender and OBI infection rate. In this study, the liver function indexes of OBI and positive control group were detected, and no significant difference was found between the two groups, indicating that the liver function indexes of OBI and hepatitis B patients were not abnormal.

The level of HBV DNA in OBI was low. In this study, HBV DNA in OBI was found to be lower than 200IU/mL. The results are similar to those of previous studies [17,18]. There was no correlation between different serum types and HBV DNA viral load levels. The viral load levels of anti-HBE + and anti-HBC + modes were slightly higher, suggesting that detection of anti-HBE + and anti-HBC + might improve the diagnosis rate of OBI.

At present, the formation mechanism of OBI has not been clearly explained, and previous reports may think that it is related to the amino acid mutation of the S gene in the coding region of HBsAg [19]. It is believed that the hydrophilic region in the S region directly affects HBsAg, and the mutation or replacement of the S location may cause antigenic changes and thus the expression of HBsAg. It can be considered that the gene mutation in this region will no longer produce the effect of neutralizing antibodies in the body, resulting in the production of OBI [20, 21].

This study found that there are B genotype and C genotype, and the mutation frequency of B genotype is higher than that of C genotype, which may be related to regional differences [22]. In this study, the number of G145R mutations in B genotype was the highest, and G145R could weaken the antigenicity of HBsAg, thereby reducing the expression of HBsAg, which had a great interference in the diagnosis of OBI [23]. There are other speculations about the mechanism of G145R, such as the possibility that it may affect the expression of HBV virus levels, resulting in lower virus levels. This study can continue to further explore the mechanism of this aspect.

Most of the gene mutation sites in this study were reported in previous literatures [24-35], such as M133L, A159V, etc., which again verified the gene mutation characteristics and formation mechanism of OBI. This study found that there is a certain correlation between HBV S gene mutation and the pathogenesis of OBI, and if the S gene mutation is likely to lead to the omission of OBI. Subsequent studies can explore from this aspect how various gene mutations lead to reduced expression of HBsAg. So it can provide scientific basis for clinical diagnosis and detection.

This study mainly studied the correlation between S region gene mutation and OBI, and made a preliminary exploration of B genotype and C genotype. According to relevant reports, there are other mutation-prone regions in OBI, such as the transmembrane structure TM, most of the amino acids in this region are hydrophobic, and the mutation of TM may affect the topology of the peptide chain of S protein. In addition, the change of hydrophobic structure may also lead to the change of antigen conformation of HBsAg, which will interfere with the detection of HBsAg. The follow-up study also made further exploration from this aspect to understand the mechanism of action of TM region and S region on OBI.

## Data Availability

All data produced in the present study are available upon reasonable request to the authors

